# Using clinical text to refine unspecific condition codes in Dutch general practitioner EHR data

**DOI:** 10.1101/2024.01.04.24300823

**Authors:** Tom M Seinen, Jan A Kors, Erik M van Mulligen, Egill Fridgeirsson, Katia MC Verhamme, Peter R Rijnbeek

## Abstract

**Objective:** Observational studies using electronic health record (EHR) databases often face challenges due to unspecific clinical codes that can obscure detailed medical information, hindering precise data analysis. In this study, we aimed to assess the feasibility of refining these unspecific condition codes into more specific codes in a Dutch general practitioner (GP) EHR database by leveraging the available clinical free text.

**Methods:** We utilized three approaches for text classification—search queries, semi-supervised learning, and supervised learning—to improve the specificity of ten unspecific International Classification of Primary Care (ICPC-1) codes. Two text representations and three machine learning algorithms were evaluated for the (semi-)supervised models. Additionally, we measured the improvement achieved by the refinement process on all code occurrences in the database.

**Results:** The classification models performed well for most codes. In general, no single classification approach consistently outperformed the others. However, there were variations in the relative performance of the classification approaches within each code and in the use of different text representations and machine learning algorithms. Class imbalance and limited training data affected the performance of the (semi-)supervised models, yet the simple search queries remained particularly effective. Ultimately, the developed models improved the specificity of over half of all the unspecific code occurrences in the database.

**Conclusions:** Our findings show the feasibility of using information from clinical text to improve the specificity of unspecific condition codes in observational healthcare databases, even with a limited range of machine-learning techniques and modest annotated training sets. Future work could investigate transfer learning, integration of structured data, alternative semi-supervised methods, and validation of models across healthcare settings. The improved level of detail enriches the interpretation of medical information and can benefit observational research and patient care.

## Introduction

Observational studies leveraging electronic health record (EHR) databases are increasingly common. These studies primarily rely on clinical codes assigned to patient records to document medical conditions and treatments. The precision of these codes is influenced by factors such as the healthcare professional’s coding skills and the terminology systems implemented within the EHR. A significant challenge arises when codes lack specificity, such as “Fracture: hand/foot bone” or “Infection of circulatory system.” These broad categories obscure the details of individual conditions, complicating the extraction of specific data like hand fractures or endocarditis cases. Consequently, this lack of detail hampers in-depth analysis.

Refining these unspecific clinical codes into more specific codes using additional clinical information can enhance observational research. However, the manual review of extensive clinical documentation to achieve this is labor-intensive and impractical for large datasets. Hence, automated classification methods that can discern the correct specific codes from clinical texts would be a substantial aid.

Automatically assigning diagnostic codes to medical texts using supervised learning algorithms has been a large focus of clinical machine learning and natural language processing (NLP) research [1,2]. Studies have been conducted in various languages [3–5], including Dutch [6–8], and have shown increasingly promising results [9–11]. These studies have primarily focused on databases or datasets containing text and labeled data, for example, codes from the International Classification of Diseases (ICD). The issue we face involves observational data with unspecific codes not already labeled with a more specific code, which precludes using a supervised approach without prior manual annotation.

When labeled data are scarce, semi-supervised or unsupervised learning methods become valuable [12]. Semi-supervised learning combines labeled and unlabelled data or employs a partial labeling process to minimize manual effort while providing results comparable to fully supervised methods [13–15]. This approach has been effectively used in clinical contexts, such as cancer risk identification [16] and biomedical text classification [17]. Unsupervised methods, including rule-based systems [18], methods relying on named entity recognition and summarization [19], and methods using word embedding similarities [20], have been applied to clinical code assignment, as well as in Dutch clinical settings [21].To our knowledge, previous research has not explored the reclassification of unspecific clinical codes into more specific ones in unlabeled EHR datasets.

In this study, we aimed to enhance the specificity of clinical codes without relying heavily on manual annotation. We evaluated and compared three classification approaches (search queries, semi-supervised learning, and supervised learning) on a selection of unspecific condition codes in a Dutch general practitioner (GP) EHR database, and determined the total number of code refinements by applying the classification models to the entire database.

## Materials and methods

### Database and population

This study used data from the Integrated Primary Care Information (IPCI) database [22], a longitudinal EHR database. IPCI covers patients from 1993 to 2022, currently holding EHRs of 2.5 million patients with a median follow-up duration of 4.8 years. The data are standardized using the Observational Medical Outcomes Partnership Common Data Model (OMOP CDM), allowing standardized analytics and facilitating international research collaboration [23–25]. The IPCI governance board approved this research under code 2022-04. All patients in the database were eligible for inclusion.

### Coded conditions

Dutch GPs code conditions using the International Classification of Primary Care (ICPC-1), a terminology containing many unspecific codes. The concept mapping from ICPC-1 to the Systematized Nomenclature of Medicine Clinical Terms (SNOMED CT), provided by the Dutch National Institute for Health Information and Communication Technology (Nictiz) ^1^, was used as starting point to identify 178 unspecific ICPC codes, defined as codes with a mapping to two or more SNOMED CT codes. For example, ICPC-1 code L74: “Fracture: hand/foot bone” maps to SNOMED CT codes 20511007: “Fracture of hand” and 15574005: “Fracture of foot”. We selected eight unspecific ICPC-1 codes from the Nictiz mapping for this feasibility study. Additionally, we identified two other ICPC-1 codes with a general or broad description not included in the mapping, D75: “Malignant neoplasm of colon/rectum” and K70: “Infection of circulatory system”. Table 1 lists the studied ICPC-1 codes and the more specific SNOMED CT codes.

**Table 1.**
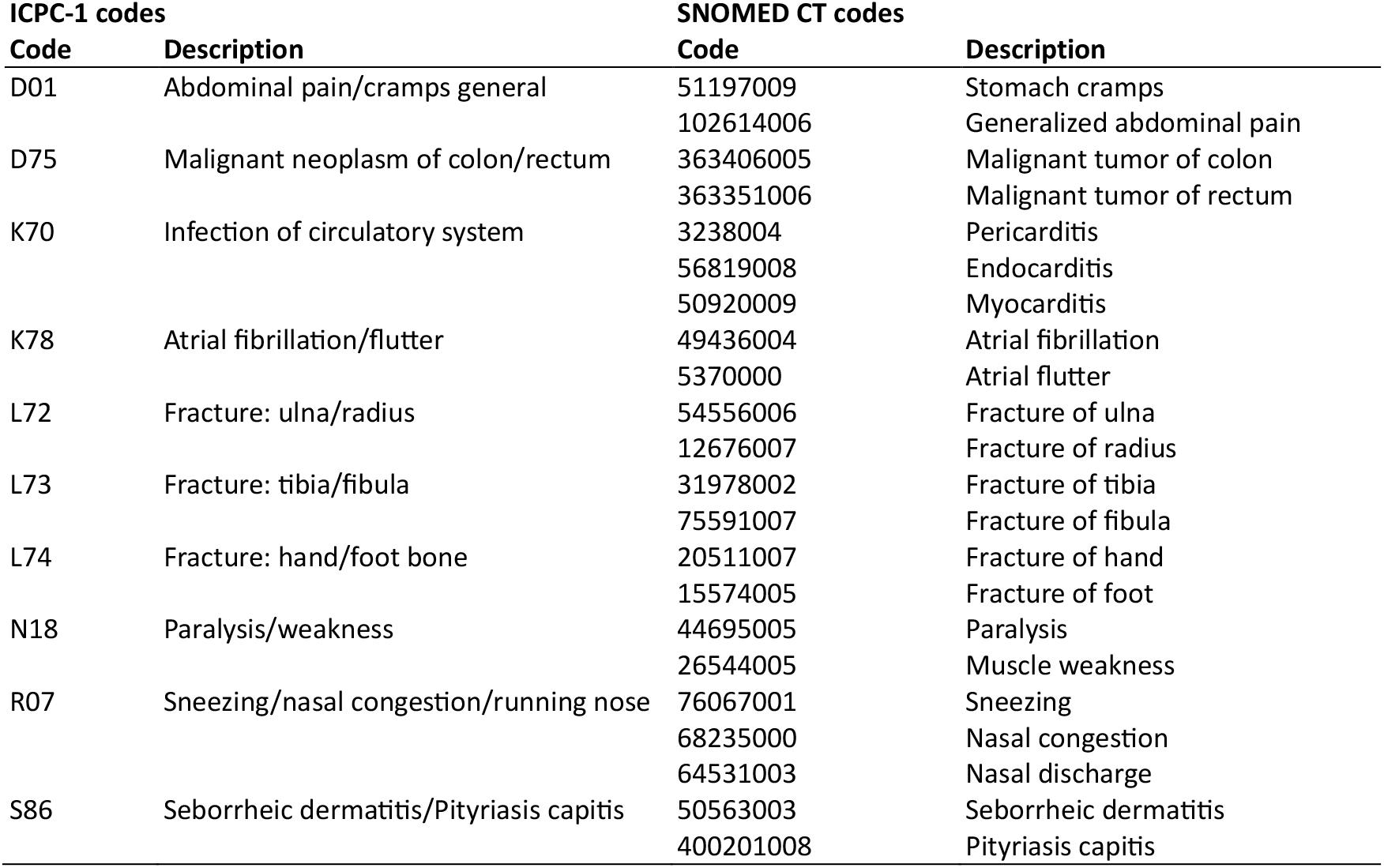
The ten unspecific ICPC-1 codes in this feasibility study with their more specific SNOMED CT codes.

### Data extraction and annotation

We extracted every occurrence of the ten selected ICPC-1 codes, along with the free-text notes within a five-day window surrounding the date each code was recorded. We randomly selected a test set of 200 code occurrences for each ICPC-1 code from the complete set of code occurrences. The first author manually annotated the code occurrences with the more specific codes, in the following also called subcodes, based on the patient’s free-text notes in the five-day window. Each subcode received a binary label, which allowed to indicate the presence of multiple subcodes, e.g., a patient could have both a broken hand and foot. A separate set of 300 randomly selected code occurrences, not in the test set, was also annotated as a modest training set for the supervised classification models. Figure 1 illustrates the experimental setup.

**Figure 1.**
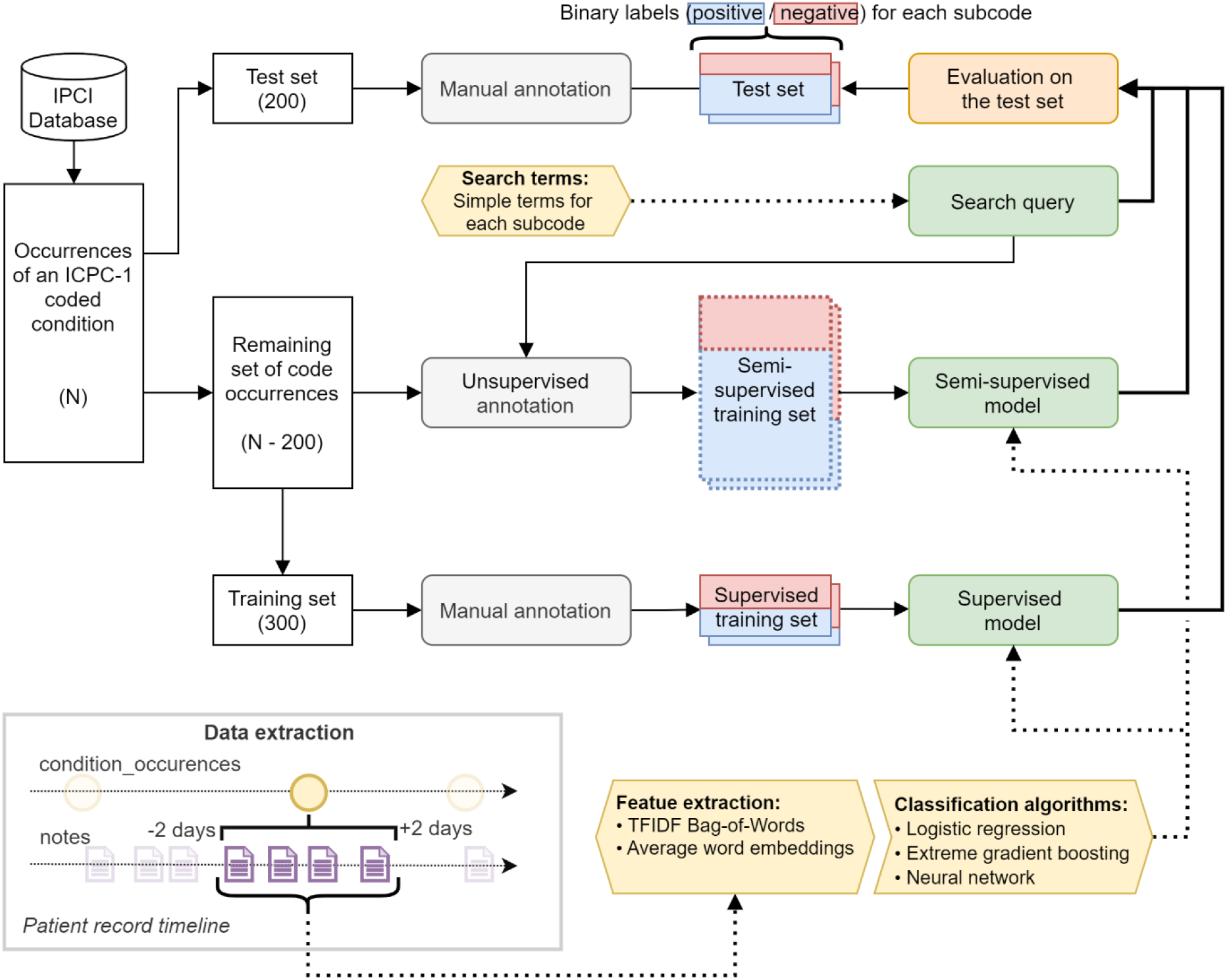
Experimental setup for the development and evaluation of classifiers that assign more specific SNOMED CT codes to unspecific ICPC-1 codes. The diagram provides an overview of the entire process, starting from the set of N code occurrences in the IPCI database and the subsequent sampling and creation of (pseudo-)labeled occurrences. The diagram also highlights the three classification approaches (search query, semi-supervised, and supervised classification) and their evaluation on the test sets. Additionally, the data extraction process is visualized.

### Classification approaches

We compared three classification approaches: search queries, semi-supervised learning, and supervised learning. In all approaches, we built a binary classifier per subcode to allow the assignment of multiple subcodes per code occurrence. For the search-query approach, we determined simple search terms for each subcode solely based on the description of the ICPC-1 code without inspection of the notes in the database. The exact substrings are in Supplemental Table S1. If we found a substring in the text, we considered the subcode present. The search queries could directly be evaluated on the test set. For the semi-supervised approach, we used self-training [15]. The search queries were used to predict the labels of all code occurrences except those in the test set. These pseudo-labels were used to train the semi-supervised classification models. For computational efficiency, we limited the training set to 150,000 occurrences. The supervised models were trained using the 300 occurrences in the manually annotated training set.

### Feature extraction

We combined and processed all text within the five-day time window around each code occurrence to create a Bag-of-Words (BoW) feature vector. Processing included conversion to lowercase and tokenization to unigrams and bigrams. We utilized two text representations: a BoW representation normalized using Term Frequency-Inverse Document Frequency (TFIDF), and averaged word embeddings (AVGEMB), allowing us to capture lexical and semantic information. We created the word embeddings by training a 300-dimensional Word2Vec model [26] using *gensim* [27], with a token window of five, on the entire IPCI database, consisting of 662 million clinical notes with 8.5 billion tokens.

### Machine learning algorithms

We used three classification algorithms for (semi-)supervised model training: L1-regularized logistic regression (LR) using the *glmnet* R-package, extreme gradient boosting (XGB) using the *xgboost* R-package, and a neural network (NN) using the *nnet* and *caret* R-package. Hyperparameters were optimized with 3-fold cross-validation (See Supplemental Table S2). Combined with the two text representations, we evaluated six different method combinations.

### Model evaluation

For each subcode, we evaluated the performance of the search query, with binary predictions, and the (semi-)supervised models, with probability scores, on the annotated test set using the area under the receiver operator characteristic curve (AUROC), the area under the precision-recall curve (AUPRC), and the F1-score for the search query and the maximum F1-score across all probability thresholds for (semi-)supervised models. Additionally, we assessed the global explainability in the LR and XGB models using TFIDF features by identifying the most important features. For LR, we measured feature importance based on the magnitude of coefficients (beta values). In XGB, we assessed feature importance using average gain, indicating each feature’s contribution to model performance. Lastly, we applied the models to the full set of occurrences (except the test set) to determine the percentage of code occurrences that were assigned at least one subcode. Here, the maximum F1-score on the test set determined the probability threshold for the (semi-)supervised models.

## Results

### Dataset characteristics and code counts

Table 2 presents the dataset characteristics per ICPC-1 code. A considerable variation in characteristics can be observed across the ICPC-1 codes. Table 3 displays the number of subcodes per condition code in the training and test sets, showing class imbalances for several codes. For example, in S86 (Seborrheic dermatitis/Pityriasis capitis) one subcode occurs in 78% of the test set code occurrences and the other subcode in only 9%. For some codes, such as L74 (*Fracture: hand/foot bone*), a subcode was assigned to almost all occurrences (95%), while for others, such as L73 (*Fracture: tibia/fibula*), a subcode could not be assigned in 37% of the occurrences because the information in the notes was insufficient. The relative outcome counts in the test and training sets were comparable across all condition codes.

**Table 2.** The dataset characteristics and the descriptive statistics of notes for the ten ICPC-1 codes in the IPCI database. D01: Abdominal pain/cramps general, D75: Malignant neoplasm of colon/rectum, K70: Infection of circulatory system, K78: Atrial fibrillation/flutter, L72: Fracture: ulna/radius, L73: Fracture: tibia/fibula, L74: Fracture: hand/foot bone, N18: Paralysis/weakness, R07: Sneezing/nasal congestion/running nose, S86: Seborrheic dermatitis/Pityriasis capitis.

| ICPC-1 code | D01 | D75 | K70 | K78 | L72 | L73 | L74 | N18 | R07 | S86 |
| --- | --- | --- | --- | --- | --- | --- | --- | --- | --- | --- |
| No. of codes | 418,143 | 145,405 | 9,735 | 352,190 | 61,992 | 46,835 | 42,894 | 22,804 | 63,558 | 125,060 |
| No. of patients | 179,263 | 14,213 | 2,663 | 48,138 | 34,148 | 18,039 | 26,583 | 9,388 | 34,839 | 68,850 |
| Mean no. of codes per patient | 2.3 | 10.2 | 3.7 | 7.3 | 1.8 | 2.6 | 1.6 | 2.4 | 1.8 | 1.8 |
| Median no. of codes per patient | 1 | 4 | 1 | 3 | 1 | 1 | 1 | 1 | 1 | 1 |
| Mean age | 39.6 | 70.1 | 59.3 | 73.4 | 42.6 | 48.2 | 40.1 | 59.5 | 42.3 | 45.6 |
| SD age | 24.5 | 11.8 | 18.1 | 11.3 | 28.3 | 23.1 | 22.5 | 20.8 | 24.2 | 24.0 |
| Percentage female | 67% | 47% | 34% | 48% | 64% | 59% | 50% | 50% | 49% | 55% |
| Median no. of notes per code | 6 | 3 | 3 | 4 | 5 | 4 | 5 | 5 | 4 | 3 |
| Median no. of characters per code | 471 | 363 | 307 | 320 | 514 | 299 | 508 | 416 | 361 | 245 |

**Table 3.**
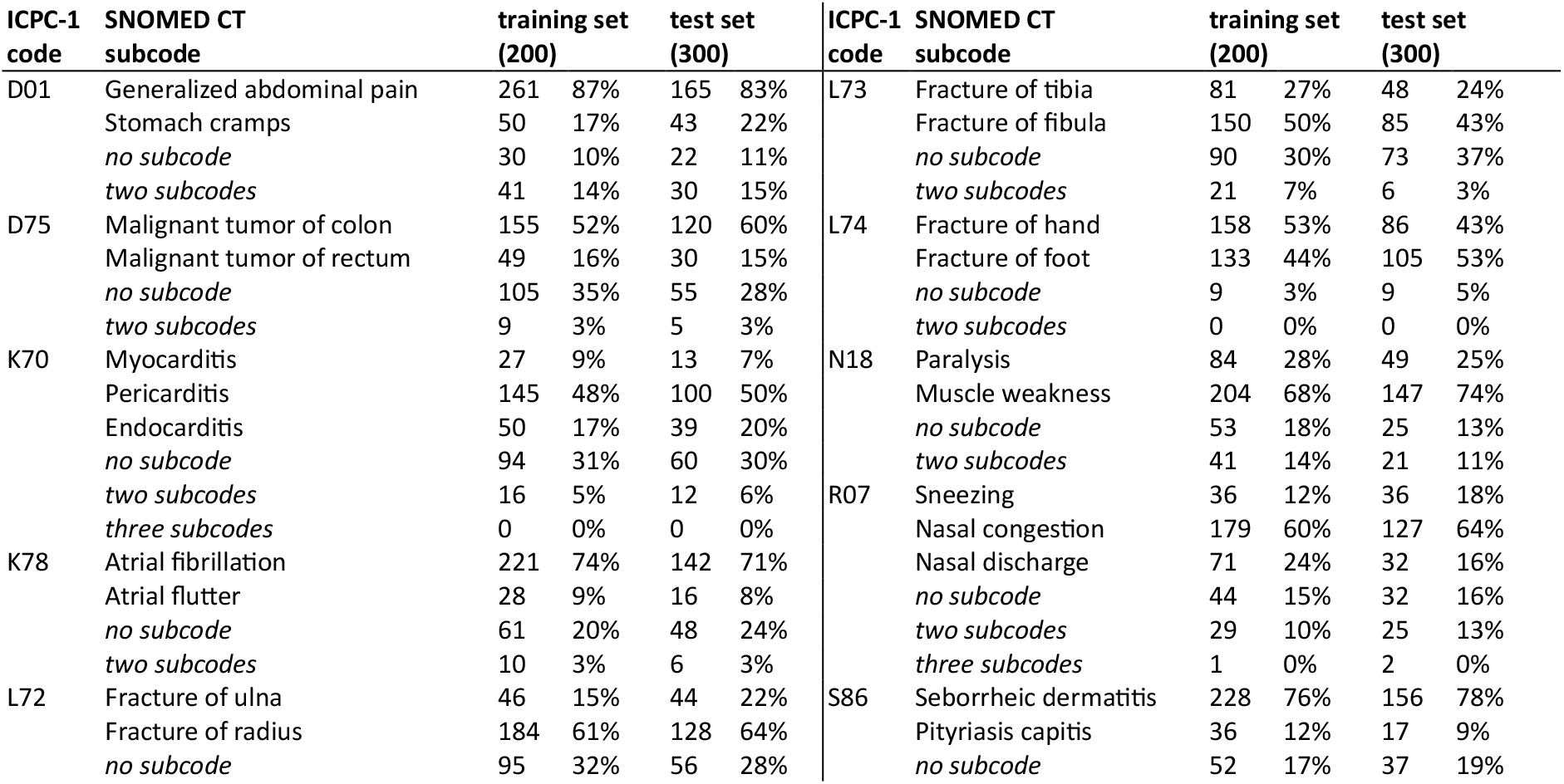
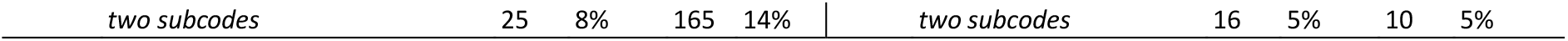
Subcode counts and percentages in the annotated test sets (200 code occurrences) and the training sets (300 code occurrences) for each of the ten ICPC-1 codes. Also shown are the number and percentage of code occurrences with no, two, and three subcodes assigned.

### Model performance

We developed 286 classification models for the 22 subcodes. For each subcode, the models comprised a search query model, six semi-supervised models, and six supervised models. Figure 2 illustrates the performance of all the models on the test sets. We observed that the models’ overall performance, ranging from good to excellent for most subcodes, was comparable across different classification approaches. However, the performance of the (semi-)supervised models varied across the different text representations and machine learning algorithms. Specifically, the supervised models, trained on the 300 annotated occurrences, achieved beter results with TFIDF features than with averaged word embeddings, especially when using XGB and LR. Semi-supervised models, trained on a larger dataset (N −200), showed a similar trend but with smaller performance differences.

**Figure 2.**
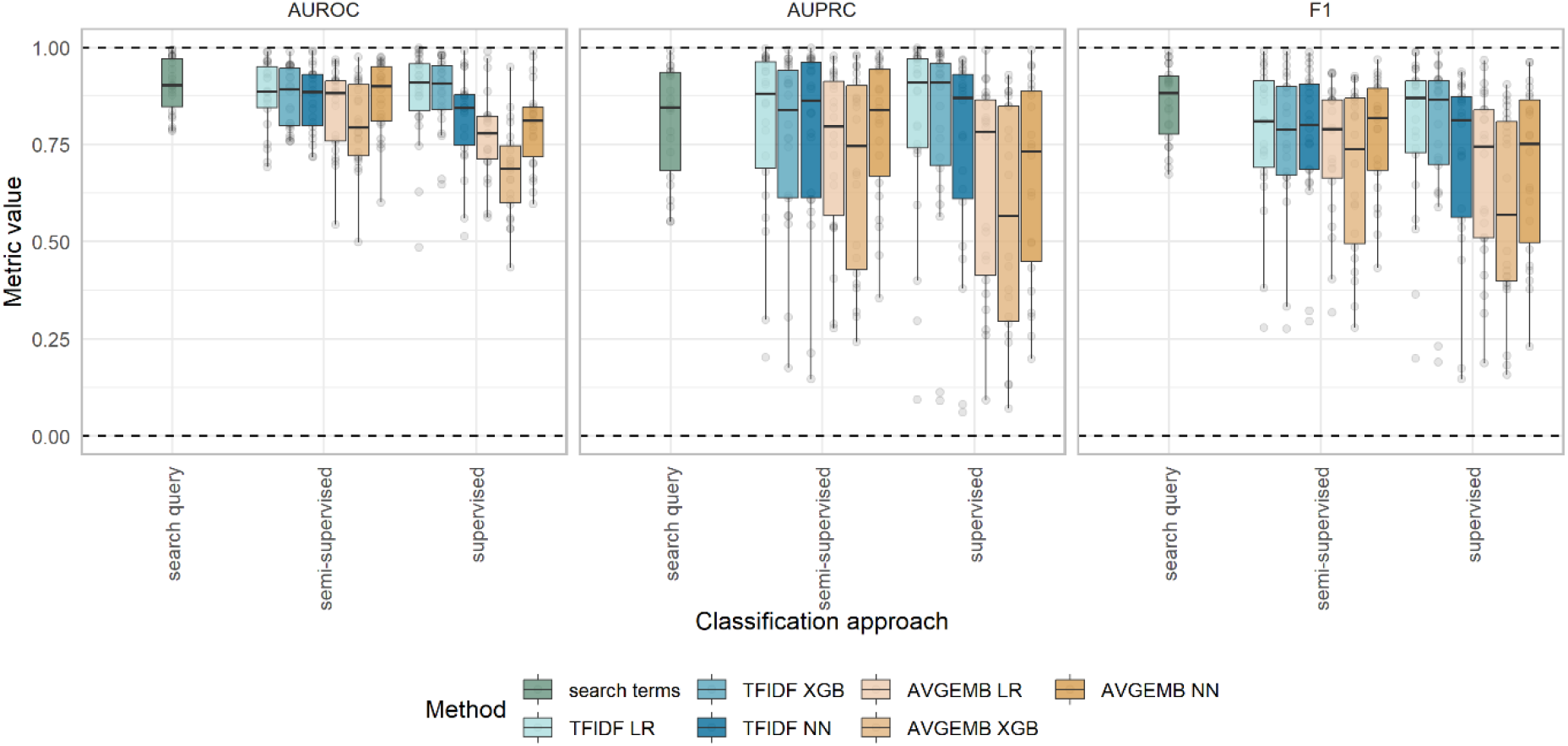
Performance on the test set across all subcode models, per classification approach (search query, semi-supervised learning, and supervised learning), method (search terms, text representation, and machine learning algorithm), and evaluation metric (AUROC, AUPRC, and F1-score). The points indicate the performance of the individual subcode models.

The average performance of the best models per classification approach was comparable, but showed notable variation across subcodes. Figure 3 presents the predictive performance of the three approaches measured by AUROC for all subcodes, with the (semi-)supervised models using LR and TFIDF features. For example, the search query model outperformed the (semi-)supervised models in classifying *atrial flutter* and *myocarditis*, while the supervised models were beter in classifying subcodes such as *fracture of hand* and *paralysis*. Supplementary Figures S1 and S2 show these same paterns for the AUPRC and F1 evaluation metrics. The complete evaluation results are available in the supplementary material.

**Figure 3.**
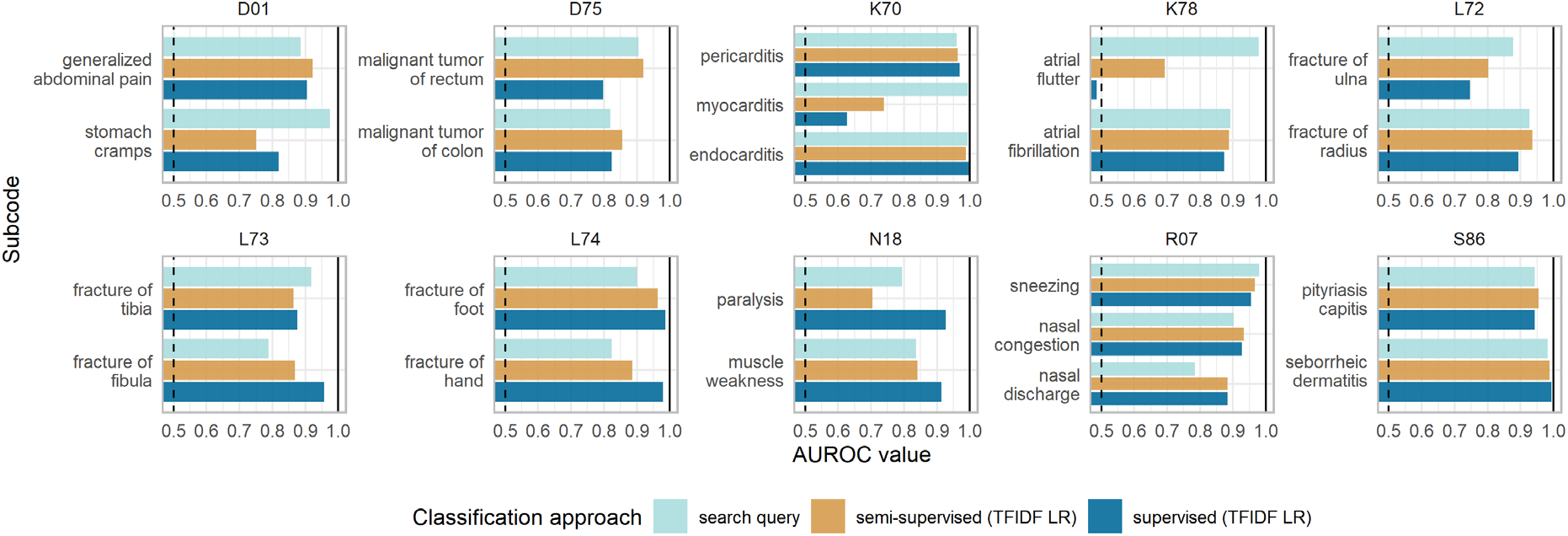
Predictive performance, measured by the AUROC, for each subcode model, developed using the search query, and semi-supervised and supervised learning using TFIDF features and regularized logistic regression, LR. The dashed line indicates an AUROC value of 0.5.

### Effect of class imbalance

To investigate the factors contributing to the suboptimal performance of several models, we examined the impact of class imbalance of subcodes on model performance. Figure 4 illustrates the relationship between the number of positive examples in the training set for the supervised models across all subcodes. As expected, subcode models with significant class imbalance, for instance, *myocarditis* and *atrial flutter*, which had fewer positive examples, tended to demonstrate poorer performance. Supplementary Figure S3 extends this analysis to the test set for all classification approaches, confirming the negative effect of class imbalance on the performance of semi-supervised models as well.

**Figure 4.**
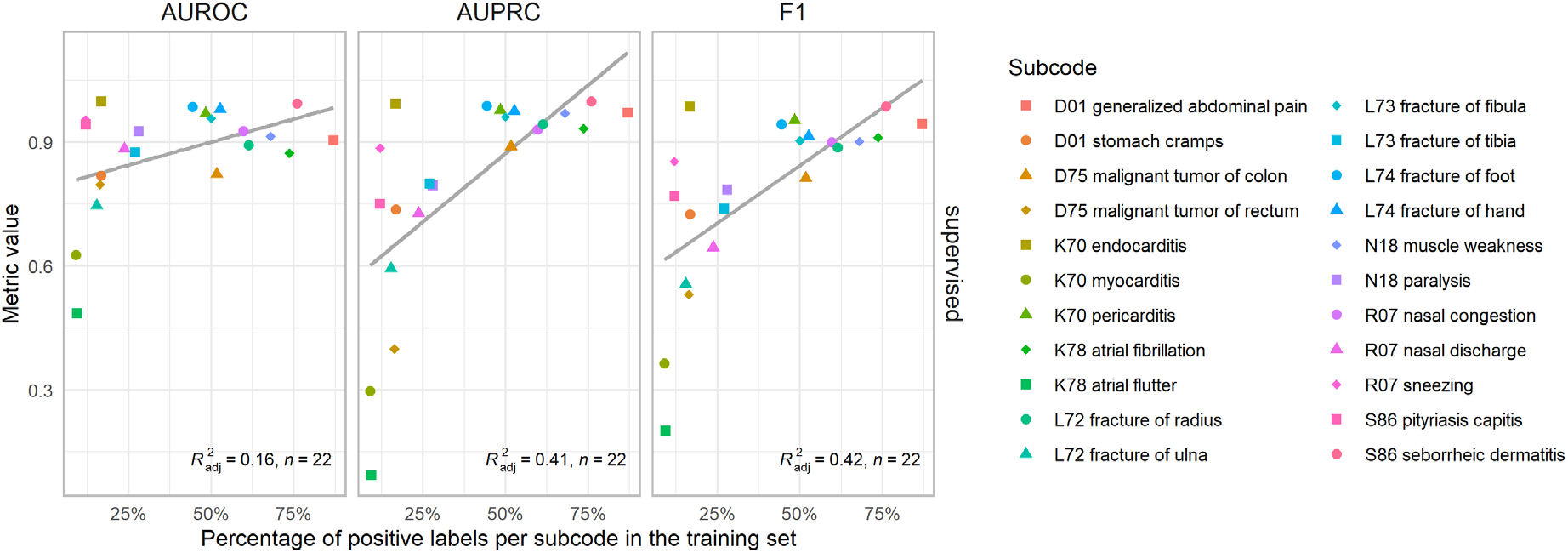
Scatterplots demonstrating the relationship between the number of positive examples in the training set in each subcode and the corresponding model performance for the supervised models using LR with TFIDF features across all subcodes. The color and shape of the points indicate the different subcodes, the grey line illustrates a regression line, and its respective adjusted R2 is presented in each graph.

### Feature importance

We investigated the most important features contributing to the model predictions to understand the performance differences between classification approaches. Table 4 lists the top-5 important features for the (semi-)supervised models (TFIDF LR) and the search query terms of three subcodes: *stomach cramps* (D01), *atrial fibrillation* (K78), and *fracture of foot* (L74). These subcodes were chosen because of their varying performances across the three classification approaches. The most important features of the other models are available in the supplementary material.

For the semi-supervised models of *stomach cramps* and *fracture of foot*, terms with high beta values were similar to the search terms. However, for *atrial flutter*, no similar term was found. Instead, terms related to the other subcode, *atrial fibrillation,* appeared with both negative and positive beta values. This likely reflects the class imbalance and the frequent co-occurrence of *atrial fibrillation* and *flutter* within the texts. The supervised model for *atrial flutter* also identified a similar *atrial fibrillation* term, but only with a negative association, yet this model was not discriminative at all. For the *stomach cramps* subcode, the supervised model, which had only two non-zero beta value terms related to the search term, surprisingly outperformed the semi-supervised model that had a greater number of related terms. For the *fracture of foot* subcode, the supervised model’s feature associations were more diverse and less focused on the term ‘foot’ than the semi-supervised model, but their performances were similar. These results and some non-intuitive findings highlight the complexity of feature importance in classification and the challenges in explaining model performance.

**Table 4.** The five features with the largest absolute non-zero beta values in the (semi-)supervised models using regularised logistic regression (LR) and TFIDF bag-of-words features for three example subcode models. The terms were translated where necessary. The search queries and the AUROC, AUPRC, and F1 score values are listed per model for ease of comparison. A term between hashtags (#) indicates an anonymization tag.

| Code: subcode | Search query<br>Term | Semi-supervised<br>Term | Beta | Supervised<br>Term | Beta |
| --- | --- | --- | --- | --- | --- |
| D01: Stomach cramps | *kramp* (cramp) | *buikkrampen* (stomach cramps) | 7.93 | *krampen* (cramps) | 2.06 |
|  |  | *def* (defecation) | 0.31 | *buikkrampen* (stomach cramps) | 1.02 |
|  |  | *diarree* (diarrhea) | 0.27 |  |  |
|  |  | *soepel* (supple) | 0.24 |  |  |
|  |  | *buikpijn* (stomach pain) | -0.28 |  |  |
| AUROC; AUPRC; F1 | 0.97; 0.84; 0.91 | 0.75; 0.56; 0.58 |  | 0.82; 0.74; 0.72 |  |
| K78: Atrial flutter | *Flutter* | *boezemfibrilleren* (atrial fibrillation) | 0.75 | *cardiologie_#hospital#*<br>(cardiology_#hospital#) | 0.62 |
|  | *fladder* (flutter) | *car* (cardiology/cardiologist) | 0.32 | *onder* (under) | 0.44 |
|  |  | *conclusie* (conclusion) | 0.30 | *mogelijk* (possible) | 0.38 |
|  |  | *af* (atrial fibrillation) | -0.25 | *huis* (home) | 0.25 |
|  |  | *atriumfibrilleren* (atrial fibrillation) | -0.40 | *atriumfibrilleren* (atrial fibrillation) | -0.58 |
| AUROC; AUPRC; F1 | 0.98; 0.67; 0.80 | 0.69; 0.20; 0.28 |  | 0.49; 0.09; 0.20 |  |
| L74: Fracture of foot | *voet* (foot) | *voet* (foot) | 17.27 | *voet* (foot) | 3.70 |
|  |  | *tijdens* (during) | 0.87 | *teen* (toe) | 1.86 |
|  |  | *lopen* (walking) | 0.53 | *kon* (could) | 1.49 |
|  |  | *mt* (metatarsal) | 0.40 | *vinger* (finger) | -1.93 |
|  |  | *teen* (toe) | 0.34 | *hand* | -2.62 |
| AUROC; AUPRC; F1 | 0.90; 0.89; 0.90 | 0.96; 0.97; 0.94 |  | 0.99; 0.99; 0.94 |  |

### Subcode assignment

To illustrate the impact of the code refinement, we applied the classification models to the set of code occurrences (N-200) for each of the ten ICPC-1 condition codes. Our study included 1,288,616 condition occurrences from 436,124 patients across all ten ICPC-1 codes. We found that 62.3% (802,931) of these occurrences could be refined to more specific codes using supervised learning (TFIDF LR). The semi-supervised method (TFIDF LR) showed a similar refinement rate of 62.2% (801,519), and the search query method reached 57.5% (740,954). Supplemental Figure S4 illustrates for each ICPC-1 code the proportion of occurrences that could be refined. Although the number of refined codes varied per ICPC-1 code, there was a high level of agreement between the different classification approaches. It should be noted that the refinement rate does not indicate the accuracy of the classifications. Instead, it highlights the potential of these models to enhance the granularity of data in the database.

## Discussion

### Condition code refinement

This study explored the feasibility of refining unspecific ICPC-1 condition codes in a Dutch GP database into more specific SNOMED subcodes using clinical free-text data. Using three different classification approaches, we developed and evaluated 286 classification models for 22 subcodes within ten different unspecific ICPC-1 codes. We found that it was possible to accurately classify each code into its subcodes using one of the different classification approaches and the information from the clinical text, proving its feasibility. In general, no single classification approach consistently outperformed the others across all subcodes. However, the relative performance of the classification approaches varied within the different subcodes.

Despite its simplicity, the search query approach using terms from the ICPC-1 code descriptions yielded good results for most subcodes. The best semi-supervised models, using TFIDF features, performed similarly. Consistent with findings from other studies, the semi-supervised models achieved results comparable to those of supervised models [13–15]. Nonetheless, inaccuracies in pseudo-labels, stemming from the choice of search terms and class imbalance within the dataset, may have negatively impacted the semi-supervised performance. The best supervised models, using TFIDF features and trained with a modest set of manually annotated code occurrences, often matched the performance of the semi-supervised and search query approaches. However, their performance was not consistent across subcodes, impacted by the class imbalance and the limited number of positive examples for certain subcodes. Regarding different text representations and machine learning algorithms, models using dense averaged word embedding features along with LR or XGB performed less well than TFIDF features, probably due to the small training set, as this effect was most pronounced in the supervised models.

Ultimately, applying the different approaches to all code occurrences, we could enhance over half of the unspecific ICPC-1 codes to a more specific SNOMED code. This significant improvement in detail underscores the clinical utility of our feasibility study and the potential of simple classification models to increase the granularity of condition coding within large-scale observational healthcare datasets.

### Strengths and limitations

Our study has several limitations. Firstly, we examined a limited number of unspecific ICPC-1 codes due to practical constraints. While we ensured a diverse range of conditions for a comprehensive overview and enhancing the generalizability of our findings, codes not included in our study may obtain different results. Secondly, we only use three classification algorithms with one lexical and semantic text representation. While more sophisticated techniques, such as deep learning models, could improve performance, they also introduce risks of overfitting [28] and complicate interpretability [29]. Our primary aim was to assess the feasibility of automatic code refinement with a manageable number of low-resource models and even with these limited methods, we were able to show a good performance for most subcodes. Thirdly, we intentionally kept our training sets small to reflect the practicality of supervised code refinement with minimal annotation efforts. However, the small size in combination with large class imbalance may have compromised the accuracy of some supervised models. Lastly, our work stands out by exploring text classification within a Dutch GP database context using three classification approaches, expanding the scope beyond the predominantly English language and hospital-based databases that have been the focus of much of the existing literature in this field [30].

### Future work

Future research in automatic code refinement could explore transfer learning with pre-trained deep learning models, such as large language models, which have shown promise in various NLP tasks and could enhance code refinement performance [9–11]. Additionally, integrating structured clinical event data and exploring other semi-supervised or ensemble methods might prove beneficial. These methods could include combining large unlabeled datasets with small labeled training sets within a single model or utilizing predictions from supervised models to inform semi-supervised learning. Expanding the focus beyond ICPC-1 to other vocabularies and validating models across different healthcare databases would be essential to assess generalizability. Automatic refinement could also facilitate the mapping of concepts between clinical terminologies, thereby enhancing the interoperability of clinical data, such as in the transition to different data models like the OMOP CDM. Lastly, conducting studies using the refined codes could illustrate their practical value in observational research, highlighting the benefits of improved data granularity in healthcare databases.

## Conclusion

In conclusion, this work successfully demonstrates that refining unspecific ICPC-1 condition codes into more specific SNOMED codes within a Dutch GP database using clinical text data and low-resource methods is feasible. We found that simple search queries were particularly effective, outperforming (semi-)supervised models when faced with issues such as class imbalance or limited training data. The enhanced granularity of coded conditions in large-scale healthcare databases could reduce manual coding costs and increase the depth and detail of data available to researchers. This improved level of detail enriches the interpretation of medical information and can benefit observational research and patient care.

## Author Statement

K.V. proposed the idea of refining unspecific ICPC-1 condition codes in the IPCI database, T.S. designed the study protocol and performed the annotations and data analysis. J.K., E.F., E.M., K.V. and P.R. provided critical feedback, helped to interpret the results and shaped the research and analysis. T.S. wrote the article with valuable input from all other authors.

## Data Availability

The aggregated data used for generating the results, conclusions, and figures/tables in this study are available as supplementary data.

## Acknowledgments

None to declare.

## Summary Table

What was already known on the topic:

- EHR databases are popular in observational research for providing large-scale data.
- The accuracy of clinical codes in these databases depends on coding proficiency and the available clinical terminologies in the EHR system.
- Unspecific concepts recorded in the EHR complicate identifying patients with specific conditions in observational research.
- Previous code classification studies focused primarily on supervised frameworks using labeled observations.

What this study added to our knowledge:

- This study successfully proved the feasibility of code refinement by classifying unspecific ICPC-1 codes to more specific SNOMED subcodes in a Dutch GP database, using code descriptions and resource-efficient classification methods supplemented by limited manual annotations.
- Three classification approaches, including search queries and semi-supervised and supervised models, generally achieved comparable results across all subcodes. However, simple search queries were especially effective in scenarios with limited training data and class imbalance.
- When applied to the entire database, the classification models could improve the specificity of more than half of the condition occurrences, indicating the potential benefit to observational research.

## Supplementary material

Appendix A: Supplementary figures and tables

Appendix B: Supplementary data

Appendix C: IJMEDI machine learning checklist

### Appendix A: Supplementary Tables and Figures

#### Supplementary Tables

**Table S1.** The terms used in the search query for each subcode. The search terms match any text containing the substrings, the sequences of characters between bars (|)

| ICPC-1 code | Dutch ICPC-1 code description | SNOMED CT Subcode | Search term |
| --- | --- | --- | --- |
| D01 | *Gegeneraliseerde buikpijn/buikkrampen* | Stomach cramps | \|pijn\| |
|  |  | Generalized abdominal pain | \|kramp\| |
| D75 | *Maligniteit colon/rectum* | Malignant tumor of colon | \|colon\| |
|  |  | Malignant tumor of rectum | \|rectum\| |
| K70 | *Infectieziekte hartvaatstelsel* | Pericarditis | \|pericarditis\| |
|  |  | Endocarditis | \|endocarditis\| |
|  |  | Myocarditis | \|myocarditis\| |
| K78 | *Boezemfibrilleren/-fladderen* | Atrial fibrillation | \|fibrill\| |
| L72 | *Fractuur radius/ulna* | Atrial flutter | \|flutter\|, \|fladder\| |
|  |  | Fracture of ulna | \|ulna\|, \|ellepijp\| |
|  |  | Fracture of radius | \|radius\|, \|spaakbeen\| |
| L73 | *Fractuur tibia/fibula* | Fracture of tibia | \|tibia\|, \|scheenbeen\| |
|  |  | Fracture of fibula | \|fibula\|, \|kuitbeen\| |
| L74 | *Fractuur hand/voet* | Fracture of hand | \|hand\| |
|  | *Verlamming/krachtverlies* | Fracture of foot | \|voet\| |
| N18 |  | Paralysis | \|verlamming\|, \|uitval\|, \|paralyse\| |
|  |  | Muscle weakness | \|krachtverlies\|, \|krachtsverlies\|, \|zwakte\|, \|parese\| |
| R07 | *Niezen/neusverstopping/loopneus* | Sneezing | \|niezen\|, \|nies\| |
|  |  | Nasal congestion | \|verstop\| |
|  |  | Nasal discharge | \|loopneus\|, \|snotneus\| |
| S86 | *Seborroïsch eczeem/roos* | Seborrheic dermatitis | \|eczeem\| |
|  |  | Pityriasis capitis | \|roos\| |

**Table S2.**
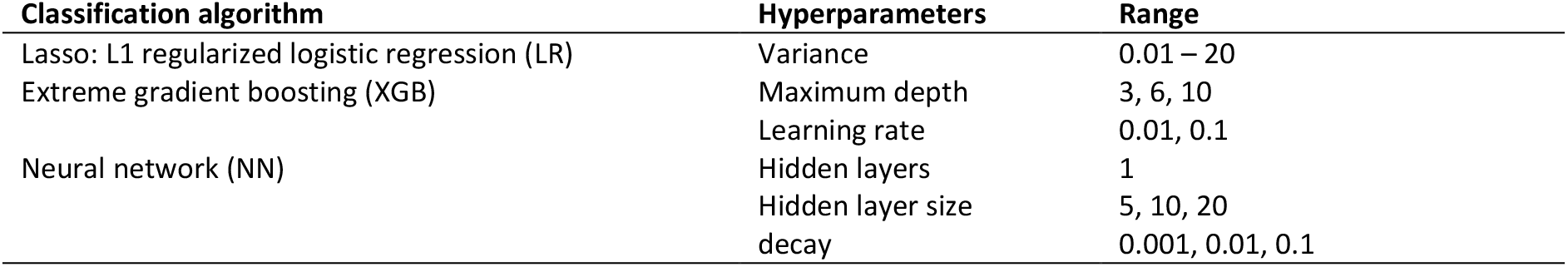
Overview of the machine learning algorithms, their hyperparameters, and hyperparameter values.

#### Supplementary Figures

**Figure S1.**
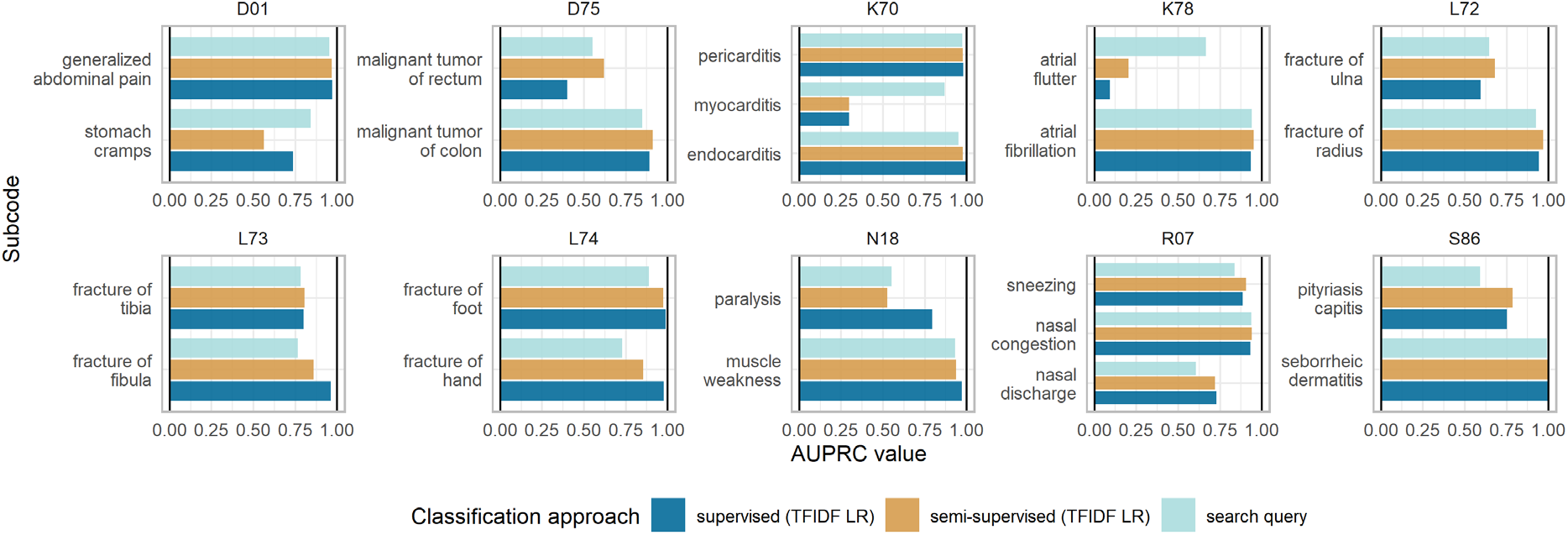
Visualization of the predictive performance, measured by the AUPRC, for each code-to- subcode model, developed using the search query and a semi-supervised and supervised model (using TFIDF features and regularized logistic regression, LR).

**Figure S2.**
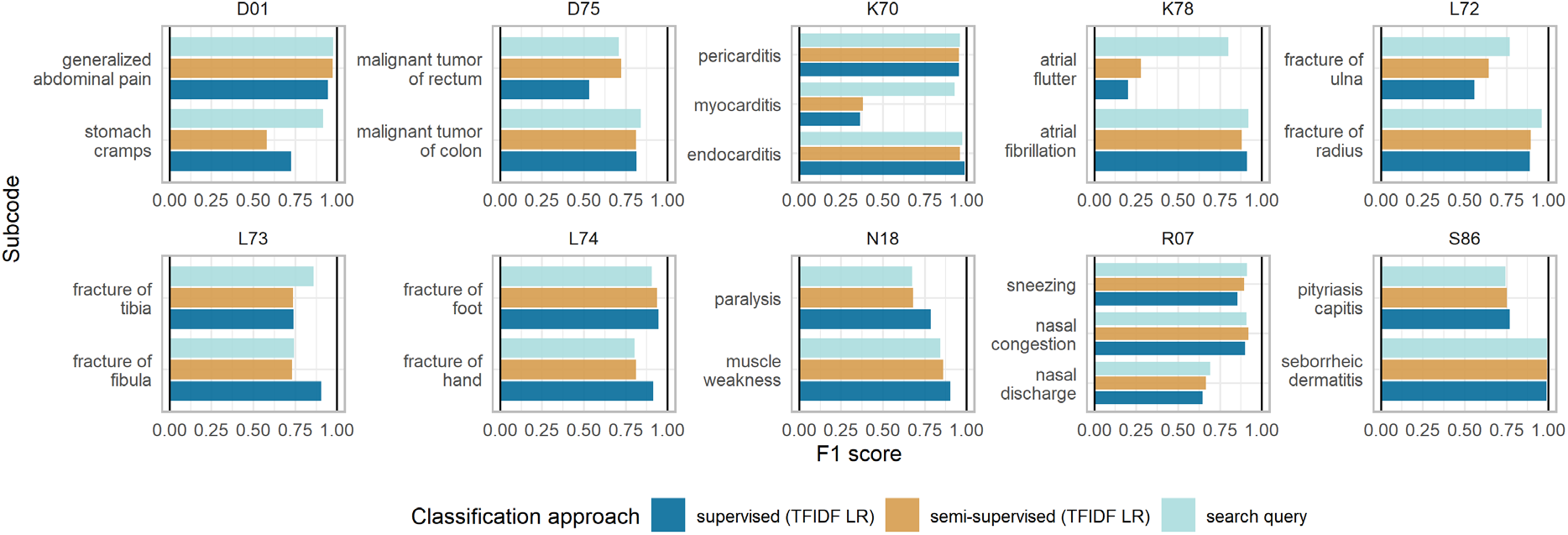
Visualization of the predictive performance, measured by the F1-score, for each code-to- subcode model, developed using the search query and a semi-supervised and supervised model (using TFIDF features and regularized logistic regression, LR).

**Figure S3.**
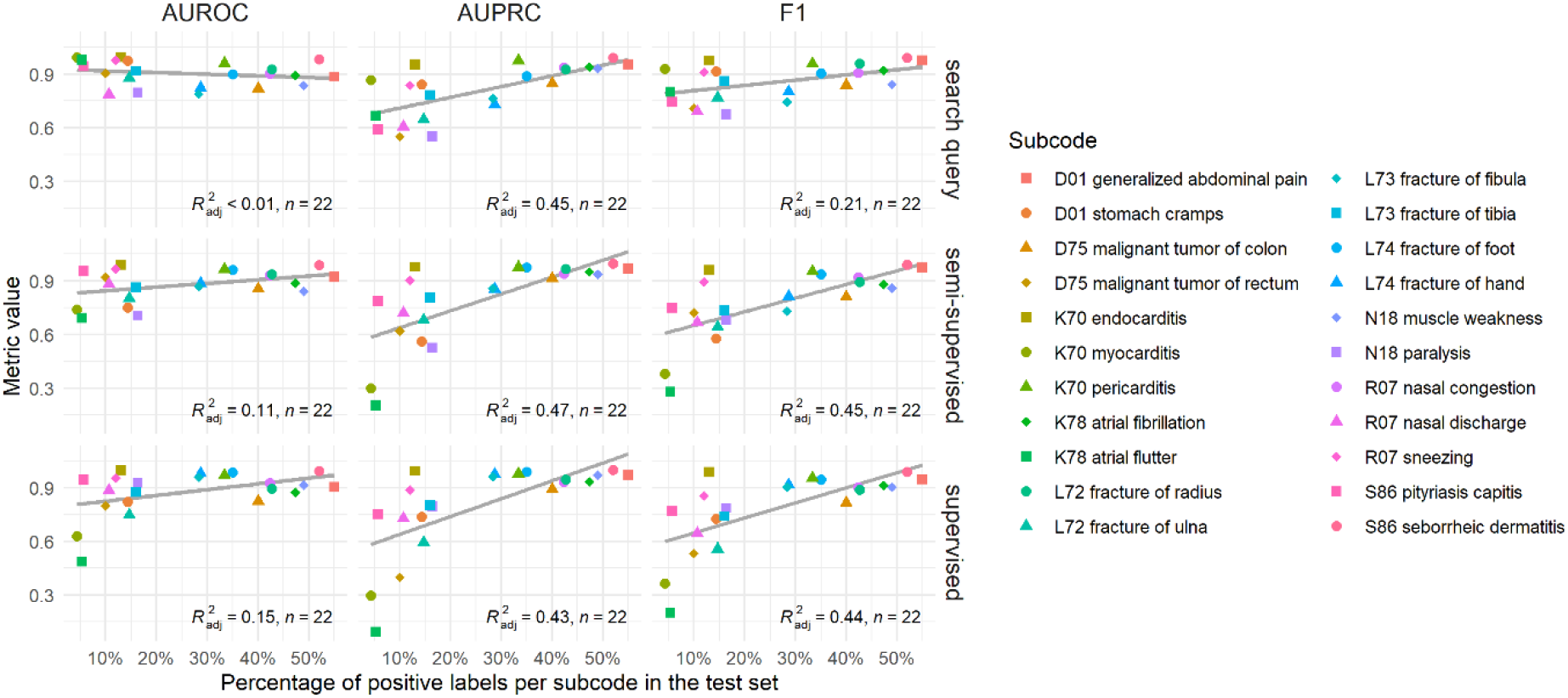
Scatterplots demonstrating the relationship between the number of positive examples in the test set in each subcode and the corresponding model performance across all subcodes and classification approaches, with the (semi-)supervised models using LR with TFIDF features. The color and shape of the points indicate the different subcodes, the grey line illustrates a regression line, and its respective adjusted R2 is presented in each graph.

**Figure S4.**
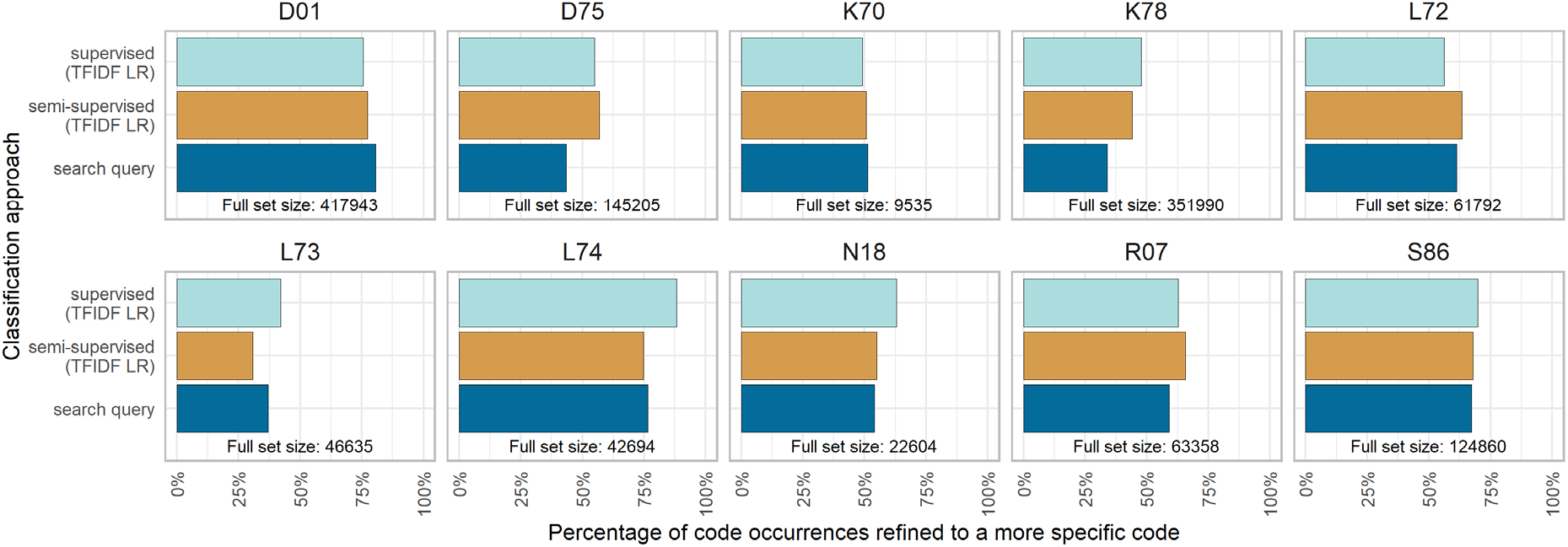
Visualization of the percentage of the code occurrences in the data set (N-200) refined to a more specific subcode per ICPC-1 code and classification approach. The semi-supervised and supervised models using LR with TFIDF features determined their threshold using the maximum F1 score. The size of the dataset is given for each ICPC-1 code.

### Appendix B: Supplementary data

#### Evaluation results

The AUROC value, AUPRC value, and F1-score for each of the 286 classification models.

CSV file: *Evaluation_results.csv*

#### Feature importance

Top 10 important features for all the LR and XGB models using TFIDF features.

CSV file: *Feature_importances.csv*

### Appendix C: IJMEDI machine learning checklist

The IJMEDI checklist for assessment of medical AI.

PDF file: *IJMEDI_ML_checklist.pdf*

## Footnotes

1 https://www.snomed.org/member/netherlands

