## Supplementary material for "Using clinical text to refine unspecific condition codes in Dutch general practitioner EHR data": IJMEDI_ML_checklist.pdf

### The IJMEDI checklist for assessment of medical AI

| Requirement | Authors | | | Reviewers | | |
|---|---|---|---|---|---|---|
| | NA | No | Yes | OK | mR | MR |
| Problem Understanding | | | | | | |
| **1. Is the study population described, also in terms of inclusion/exclusion criteria (e.g., patients older than 18 tested for COVID-19; all inpatients hospitalized for 24 or more hours)? §** | ○ | ○ | X | ○ | ○ | ○ |
| **2. Is the study design described? (e.g., retrospective, prospective, cross-sectional [1], observational, randomized control trial [2]) §** | ○ | ○ | X | ○ | ○ | ○ |
| 3. Is the study setting described? (e.g., teaching tertiary hospital; primary care ambulatory, nursing home, medical laboratory, R&D laboratory) § | ○ | ○ | X | ○ | ○ | ○ |
| 4. **Is the source of data described? (e.g., electronic specialty registry; laboratory information system; electronic health record; picture archiving and communication system) §** | ○ | ○ | X | ○ | ○ | ○ |
| **5. Is the medical task reported? (e.g., diagnostic detection, diagnostic characterization, diagnostic staging, prognosis (on which endpoint), event prediction, risk stratification, anatomical structure segmentation, treatment selection and planning, monitoring)** § | ○ | ○ | X | ○ | ○ | ○ |
| 6. Is the data collection process described, also in terms of setting-specific data collection strategies (e.g. whether body temperatures are measured only in the morning; whether some blood tests are performed only in light of a specific diagnostic hypothesis)? Any consideration about data quality is appreciated, e.g., in regard to completeness, plausibility, and robustness with respect to upcoding or downcoding practices | ○ | ○ | X | ○ | ○ | ○ |
| Data Understanding | | | | | | |
| **7. Are the subject demographics described in terms of**<br>**1. average age (mean or median);**<br>**2. age variability (standard deviation (SD) or inter-quartile range (IQR));**<br>**3. gender breakdown (e.g., 55% female, 44% male, 1% not reported); §**<br>**4. main comorbidities;**<br>**5. ethnic group (e.g., Native American, Asian, South East Asian, African, African American, Hispanic, Native Hawaiian or Other Pacific Islander, European or American White);**<br>**6. socioeconomic status?** | ○ | ○ | X | ○ | ○ | ○ |
| **8. If the task is supervised, is the gold standard described? (e.g., "100 manually annotated clinical notes and pain scores recorded in EHR, Death, re-admission and International Classification of Disease (ICD) codes in discharge letters"). In particular, the authors should describe the process of ground truthing described in terms of:**<br>**1. Number of annotators (raters) producing the labels;**<br>**2. Their profession and expertise (e.g., years from specialization or graduation);**<br>**3. Particular instructions given to annotators for quality control (e.g., which data were discarded and why);**<br>**4. Inter-rater agreement score (e.g., Alpha [3], Kappa [4], Rho [5]);**<br>**5. Labelling technique (e.g., majority voting, Delphi method [6], consensus iteration).** | ○ | ○ | X | ○ | ○ | ○ |

| Requirement | Authors | | | Reviewers | | |
|---|---|---|---|---|---|---|
| | NA | No | Yes | OK | mR | MR |
| **9. In the case of tabular data, are the features described (also in regard to how they were used in the model in terms of categories or transformation)? This description should be done for all, or, in the case that the features exceed 20, for a significant subset of the most predictive features in the following terms: name, short description, type (nominal, ordinal, continuous), and**<br>**1. If continuous: unit of measure, range (min, max), mean and standard deviation (or median and IQR). Violin plots of some relevant continuous features are appreciated. If data are hematochemical parameters, also mention the brand and model of the analyzer equipment.**<br>**2. If nominal, all codes/values and their distribution. Feature transformation (e.g. one-hot encoding) should be reported if applied. Any terminology standard should be explicitly mentioned (e.g., LOINC [7], ICD-11 [8], SNOMED [9]) if applied.** | ○ | ○ | X | ○ | ○ | ○ |
| Data Preparation | | | | | | |
| **10. Is outlier detection and analysis performed and reported? If the answer is yes, the definition of an outlier should be given and the techniques applied to manage outliers should be described (e.g., removal through application of an Isolation Forest model).** | X | ○ | ○ | ○ | ○ | ○ |
| **11. Is missing-value management described? This description should be reported in the following terms:**<br>**1. The missing rate for each feature should be reported;**<br>**2. The technique of imputation, if any, should be described, and reasons for its choice should be given. If the missing rate is higher than 10%, a reflection about the impact on the performance of a technique with respect to others would be appreciable [10].** | X | ○ | ○ | ○ | ○ | ○ |
| **12. Is feature pre-processing performed and described? This description should be reported in terms of scaling transformations (e.g. normalization, standardization, log-transformation) or discretization procedures applied to continuous features, and encoding of categorical or ordinal variables (e.g., one-hot encoding, ordinal encoding).** | ○ | ○ | X | ○ | ○ | ○ |
| **13. Is data imbalance analysis and adjustment performed and reported? The authors should describe any imbalance in the data distribution, both in regard to the target (e.g. only 10% of the patients were affected by a given disease); and in regard to important predictive features (e.g. female patients accounted for less than 10% of the total cases). The authors should also report about any technique (if any) applied to adjust the above mentioned imbalances (e.g. under- or over-sampling, SMOTE).** | ○ | ○ | X | ○ | ○ | ○ |
| Modeling | | | | | | |
| **14. Is the model task reported? (e.g., binary classification, multi-class classification, multi-label classification, ordinal regression, continuous regression, clustering, dimensionality reduction, segmentation) §** | ○ | ○ | X | ○ | ○ | ○ |
| **15. Is the model output specified? (e.g., disease positivity probability score, probability of infection within 5 days, postoperative 3-month pain scores) §** | ○ | ○ | X | ○ | ○ | ○ |
| **16. Is the model architecture or type described? (e.g., SVM, Random Forest, Boosting, Logistic Regression, Nearest Neighbors, Convolutional Neural Network)** | ○ | ○ | X | ○ | ○ | ○ |
| Validation | | | | | | |
| **17. Is the data splitting [11] described (e.g., no data splitting;, k-fold cross-validation (CV); nested k-fold CV; repeated CV; bootstrap validation; leave-one-out CV; 80%/10%10% train/validation/test)? In the case of data splitting, the authors must explicitly state that splitting was performed before any pre-processing steps (e.g. normalization, standardization, missing value imputation, feature selection) or model construction steps (training, hyper-parameter optimization), so to avoid data leakage [12] and overfitting.** | ○ | ○ | X | ○ | ○ | ○ |

| Requirement | Authors | | | Reviewers | | |
|---|---|---|---|---|---|---|
| | NA | No | Yes | OK | mR | MR |
| **18. Is the model training and selection described? In particular, the training procedure, hyper-parameter optimization or model selection should be described in terms of**<br>**1. Range of hyper-parameters [13];**<br>**2. Method used to select the best hyper-parameter configuration (e.g., Hyper-parameter selection was performed through nested k-fold CV based grid search);**<br>**3. Full specification of the hyper-parameters used to generate results [13];**<br>**4. Procedure (if any) to limit over-fitting, in particular as related to the sample size [14].** | ○ | ○ | X | ○ | ○ | ○ |
| **19. (classification models) Is the model calibration described? If the answer is yes, the Brier score should be reported, and a calibration plot should be presented [15]** | ○ | X | ○ | ○ | ○ | ○ |
| **20. Is the internal/internal-external model validation procedure described [11, 16] (e.g., internal 10-fold CV, time-based cross-validation)? The authors should explicitly specify that the sets have been splitted before normalization, standardization and imputation, to avoid data leakage [12] (also refer to item 17 of this guideline). If possible, the authors should also comment on the adequacy of the available sample size for model training and validation [17, 14]. Moreover, the authors should try to choose the test set so that it is the most diverse with respect to the remainder of the sample [18] (w.r.t. some multivariate similarity function) and how this choice relates to conservative (and lower-bound) estimates of the model's accuracy (and performance)** | ○ | ○ | X | ○ | ○ | ○ |
| 21. Has the model been externally validated [19]? If the answer is yes, the characteristics of the external validation set(s) should be described. For instance, the authors could comment about the heterogeneity of the data with respect to the training set (e.g., degree of correspondence $\Psi$ [18], Data Representativeness Criterion [20]) and the cardinality of the external sample [21]. If the performance on external datasets is found to be comparable with (or better than) that on training and internal datasets, the authors should provide some explanatory conjectures for why this happened (e.g., high heterogeneity of the training set, high homogeneity of the external dataset) | ○ | X | ○ | ○ | ○ | ○ |

| Requirement | Authors | | | Reviewers | | |
| --- | --- | --- | --- | --- | --- | --- |
| | NA | No | Yes | OK | mR | MR |
| **22. Are the main error-based metrics used?**<br><br>**1. a. Classification performance should be reported in terms of: Accuracy, Balanced accuracy, Specificity, Sensitivity (recall), Area Under the Curve (if the positive condition is extremely rare - as in case of stroke events - authors could consider the “Area under the Precision-Recall Curve” [22]). Optionally also in terms of: positive and negative predictive value, F1 score, Matthew coefficient [23], F score of sensitivity and specificity, the full confusion matrix, Hamming Loss (for multi-label classification), Jaccard Index (for multi-label classification).**<br>**2. Regression performance should be reported in terms of: $R^2$; Mean Absolute Error (MAE); Root Mean Square Error (RMSE); Ratio between MAE (or RMSE) and SD (of the target)**<br>**3. Clustering performance should be reported in terms of: External validation metrics (e.g. mutual information, purity, Rand index), when ground truth labels are available, and Internal validation metrics (e.g. Davies-Bouldin index, Silhouette index, Homogeneity). The reported results of internal validation metrics should be discussed [24]**<br>**4. Image segmentation performance, depending on the specific task, should be reported in terms of metrics like [25]: accuracy-based metrics (e.g. Pixel accuracy, Jaccard Index, Dice Coefficient), distance-based metrics (e.g. mean absolute, or maximum difference), or area-based metrics (e.g. true positive fraction, true negative fraction, false positive fraction, false negative fraction).**<br>**5. Reinforcement learning performance, depending on the specific task, should be reported in terms of metrics like [26]: Fixed-Policy Regret, Dispersion across Time, Dispersion across Runs, Risk across Time, Risk across Runs, Dispersion across Fixed-Policy Rollouts, Risk across Fixed-Policy Rollouts.**<br><br>**The above estimates should be expressed, whenever possible, with their 95% (or 90%) confidence intervals (CI), or with other indicators of variability [27], with respect to the evaluation metrics reported. In this case, the authors should report which methods were applied for the computation of the confidence intervals (e.g. whether k-fold CV or bootstrap was applied, normal approximation). When comparing multiple models, the authors should discuss the statistical significance of the observed differences [28] (e.g. through CI comparisons, or hypothesis testing). When comparing multiple regression models, a Taylor diagram [29] could be reported and discussed.** | ○ | ○ | X | ○ | ○ | ○ |
| 23. Are some relevant errors described? The authors should describe the characteristic of some noteworthy classification errors or cases for which the regression prediction was much higher ($> 2x$) than the MAE. If these cases represent statistical outliers for some covariates, the authors should comment on that. To detect relevant cases, the authors could focus on those cases on which the inter-rater agreement (either re ground truth or by comparing human vs. model’s performance) is lowest. | ○ | ○ | X | ○ | ○ | ○ |
| Deployment | | | | | | |
| 24. Is the target user indicated? (e.g., clinician, radiologist, hospital management team, insurance company, patients) § | ○ | ○ | X | ○ | ○ | ○ |
| 25. (classification models) Is the utility of the model discussed? The authors should report the performance of a baseline model (e.g., logistic regression, Naive Bayes). Additionally, the authors could report the Net Benefit [30] or similar metrics and present utility curves [31]. In particular, the authors are encouraged to discuss the selection of appropriate risk thresholds [32]; the relative value of benefits (true positives/negatives) and harms (false positives/negatives); and the clinical utility of the proposed models [14]. | ○ | ○ | X | ○ | ○ | ○ |

| Requirement | Authors | | | Reviewers | | |
|---|---|---|---|---|---|---|
| | NA | No | Yes | OK | mR | MR |
| 26. Is information regarding model interpretability and explainability available [33] (e.g. feature importance, interpretable surrogate models, information about the model parameters)? Claims of "high" or "adequate" model interpretability (e.g., by means of visual aids like decision trees, Variable Importance Plots or Shapley Additive Exlanations Plots (SHAP) [34]) or model causability [35] should always be supported by some user study, even qualitative or questionnaire-based [36]. In the case surrogate models were applied, the authors should report about their fidelity [37, 38] | ○ | ○ | X | ○ | ○ | ○ |
| 27. Is there any discussion regarding model fairness, ethical concerns or risks of bias [14, 39] (for a list of clinically relevant biases, refer to [40] )? If possible, the authors should report the model performance stratified for particularly relevant population strata [41] (e.g. model performance on male vs female subjects, or on minority groups) | ○ | X | ○ | ○ | ○ | ○ |
| 28. Is any point made about the environmental sustainability of the model, or about the carbon footprint [42], of either the training phase or inference phase (use) of the model? If the answer is yes, then such a footprint should be expressed in terms of carbon dioxide equivalent ($CO_2$eq) and details about the estimation method should be given. Any efforts to this end will be appreciated, including those based on tools available online[1], as well as any attempts to popularise this concept, e.g. through equivalences with the consumption of everyday devices such as smartphones or kilometres travelled by a fossil-fuelled car[2] | ○ | ○ | X | ○ | ○ | ○ |
| **29. Is code and data shared with the community [13, 43]? § If not, are reasons given? If code and data are shared, institutional repositories such as Zenodo should be preferred to private-owned repositories (arxiv, GitHub). If code is shared, specification of dependencies should be reported and a clear distinction between training code and evaluation code should be made. The authors should also state whether the developed system, either as a sand-box or as fully-operating system, has been made freely accessible on the Web.** | ○ | X | ○ | ○ | ○ | ○ |
| 30. Is the system already adopted in daily practice? If the answer is *yes*, the authors should report on where (setting name) and since when. Moreover, appreciated additions would regard: the description on the digitized workflow integrating the system; any comment about the level of use [14]; a qualitative assessment of the level of efficacy of the system's contribution to the clinical process (e.g., [44, 45]); any comment about the technical and staff training effort actually required [14]. If the answer is *no*, the authors should be explicit in regard to the point in the clinical workflow where the ML model should be applied, possibly using standard notation (e.g., BPMN). Moreover, the authors should also propose an assessment of the technology readiness of the described system, with explicit reference to the Technology Readiness Level framework[3] or to any adaptation of this framework to the AI/ML domain [46]. In either above cases (yes/no), the authors should report about the procedures (if any) for performance monitoring, model maintenance and updating [47]. | ○ | X | ○ | ○ | ○ | ○ |

Table 1: Checklist for assessment of requirements and recommendations for sound medical ML contributions to the existing literature. NA: not applicable; mR: minor revisions needed; MR: major revisions needed. Items in bold indicate priority aspects to be considered. Items denoted with a § symbol are directly inspired by the MINIMAR guideline [48]. The section names for the checklist items are directly inspired by the CRISP-DM framework [49].

[1] https://mlco2.github.io/impact/

[2] https://www.epa.gov/energy/greenhouse-gas-equivalencies-calculator

[3] Technology readiness levels (TRL) - Extract from Part 19 - Commission Decision C (2014) 4995

If you want to cite this checklist: Cabitza F., Campagner, A. (2021) The need to separate the wheat from the chaff in medical informatics. International Journal of Medical Informatics.

This work is licensed under a Creative Commons "Attribution-ShareAlike 4.0 International" license.
